# Decoupling Reasoning and Reward: A Modular Approach for Stable Alignment of Small Clinical Language Models

**DOI:** 10.64898/2026.03.12.26348283

**Authors:** Kiran Bhattacharyya, Sreeram Kamabattula

## Abstract

Deploying language models (LMs) in clinical settings requires navigating competing demands between accuracy, auditability, and on-device efficiency for privacy. While smaller LMs are desirable for this purpose, aligning them with methods like Group Relative Policy Optimization (GRPO) is often hindered by training instability and objective conflicts. Prior work has shown that Chain-of-Thought (CoT) supervision during supervised fine-tuning (SFT) can stabilize GRPO, but existing approaches typically entangle these objectives within a single, monolithic model. In this work, we introduce a modular, adapter-based alignment framework that decouples reasoning supervision and reward tuning into separate, composable parameter-efficient fine-tuning (PEFT) stages using LoRA adapters. We evaluate five alignment configurations on a medically grounded question answering dataset, using Qwen2.5 models from 0.5B to 7B to analyze how alignment stability, factual accuracy, and structural auditability scale with model size. Our findings demonstrate that this modular approach resolves key training instabilities, especially in smaller models, and produces structurally consistent, auditable reasoning without sacrificing accuracy. To support further research, we release, upon publication, (1) our dataset comprising over 100K clinically relevant QA pairs with CoT traces and (2) our multi-stage alignment codebase. We conclude that decoupling reasoning and reward offers a flexible and robust foundation for building privacy-preserving, verifiably aligned clinical LLMs that successfully address the competing demands in clinical AI.

## 1 Introduction

Large language models (LLMs) have demonstrated strong performance on clinical reasoning and question answering tasks. However, deploying these models in *safety-critical settings like healthcare* remains challenging due to a fundamental tension between 3 competing demands—1)*accuracy and reliability* in providing evidence-based answer; 2)*verifiability and auditability* ensuring transparent and structurally predictable reasoning; and 3) *efficiency and privacy*. In contexts such as point-of-care tools or embedded clinical systems, models must run behind institutional firewalls or on edge devices with limited resources Jonnagaddala and Wong [2025], Garg et al. [2025]. While smaller LMs are an appealing solution to the efficiency and privacy challenge, they often struggle to meet the demands for accuracy and auditability, particularly when subjected to standard alignment procedures Dang and Ngo [2025].

Alignment methods using reward signals from humans or verifiable criteria such as Reinforcement Learning with Human Feedback (RLHF) Ouyang et al. [2022] and Group Relative Policy Optimization (GRPO) Shao et al. [2024] are effective for tuning model factuality, but they often suffer from instability and objective conflicts when applied to smaller models Dang and Ngo [2025], compromising convergence. This instability is a critical barrier, as it degrades both accuracy and the structural consistency in reasoning required for auditability. Recent work Guo et al. [2025] shows that supervising models with *Chain-of-Thought (CoT)* traces during supervised fine-tuning (SFT) significantly improves the stability and performance of subsequent GRPO alignment. Yet, existing implementations for medical reasoning Chen et al. [2024] typically entangle reasoning and reward signals into a monolithic model, limiting adaptability and modularity.

In this work, we propose a new alignment framework that decouples reasoning supervision and reward tuning into *modular, independently trained adapters* using parameter-efficient fine-tuning (PEFT) via LoRA Hu et al. [2022]. By separating SFT and GRPO into distinct adapters, we enable composability, adapter reuse, and targeted debugging—features that are especially beneficial in constrained medical contexts where explainability and auditability are essential.

We systematically compare five alignment configurations:

1. Base model (instruction fine-tuned),
2. SFT with CoT traces from base model using LoRA,
3. GRPO from base model with LoRA,
4. SFT adapter frozen, followed by GRPO adapter training,
5. Unified adapter trained sequentially with both SFT then GRPO.

Experiments are conducted on a medically grounded QA dataset that includes verifiable answer keys. We evaluate across Qwen2.5 models from 0.5B to 7B Qwen et al. [2025], analyzing how alignment strategies perform across model sizes and out-of-domain datasets. Our results confirm prior findings that CoT guidance improves GRPO training, but further demonstrate that *modular alignment* yields comparable or superior performance, with added benefits for adapter fusion and transfer.

### Our contributions are

- We introduce a modular PEFT training pipeline that separates reasoning and reward adapters, enhancing training stability and structural auditability.
- We benchmark five alignment configurations on clinical QA across model sizes, demonstrating the benefits of decoupling for smaller models.
- We provide a structured dataset of over 100K medically grounded QA pairs with CoT traces from multiple models of varying complexity to facilitate reproducible alignment research.

## 2 Background and Related Work

### 2.1 Modular Alignment with Chain-of-Thought and Reward Optimization

Large language models (LLMs) are commonly aligned using multi-stage pipelines that begin with supervised fine-tuning (SFT) on instruction-following data, followed by reward-based optimization such as reinforcement learning with human feedback (RLHF), direct preference optimization (DPO) Rafailov et al. [2023], or group relative policy optimization (GRPO) Shao et al. [2024]. GRPO has gained traction as a lightweight and stable alternative to RLHF, especially in domains requiring verifiable outputs and hand-crafted reward structures Guo et al. [2025].

However, reward-based methods can be unstable when applied directly to small or mid-sized models Xu et al. [2025], Dang and Ngo [2025], particularly in complex or data-scarce domains. Recent work has shown that intermediate reasoning supervision—especially via Chain-of-Thought (CoT) prompting—can stabilize alignment and improve generalization Wei et al. [2022]. While early CoT approaches focused on zero-shot prompting, fine-tuning with CoT traces has been shown to improve factual accuracy and verifiability, especially in high-stakes reasoning tasks Lobo et al. [2024], Li et al. [2025].

Despite these benefits, most prior work integrates CoT supervision and reward optimization within a single model Luong et al. [2024], Chen et al. [2024], limiting transparency and adaptability. We instead adopt a modular approach using parameter-efficient fine-tuning (PEFT) with LoRA Hu et al. [2022], which allows us to decouple the reasoning and reward components into separate adapters. This modularity supports independent training, auditing, and composition—features particularly useful in clinical or high-risk deployment settings. While PEFT methods have seen widespread use in medical LLMs Gema et al. [2023], Han et al. [2023], Christophe et al. [2024], their potential for structured, multi-stage alignment has not been systematically explored.

Our work fills this gap by evaluating modular CoT and GRPO adapters across scales and domains, highlighting the benefits of adapter-based composability for small, verifiable, and trustworthy LLM alignment. We also introduce and implement a fuzzy-match accuracy reward for GRPO which leads to stable model training and performance, while previous work uses an exact match reward Shao et al. [2024], Guo et al. [2025].

### 2.2 Medical Question Answering and Clinical LLMs

Medical question answering (QA) is a challenging benchmark for language models due to its reliance on domain-specific knowledge, clinical reasoning, and verifiability. Datasets such as MedQA Jin et al. [2021] and MedMCQA Pal et al. [2022] have been widely used to evaluate LLMs in the biomedical domain. However, prior work often focuses on maximizing performance on in-domain benchmarks without examining alignment stability, verifiability, or deployment constraints Zhang et al. [2025], Chen et al. [2024].

Recent instruction-tuned medical models such as HuatuoGPT Zhang et al. [2023a], Med-Alpaca Han et al. [2023], and AlpaCare Zhang et al. [2023b] have demonstrated the potential of open-source LLMs for clinical QA. Moreover, the MedGemma model suite Sellergren et al. [2025] was also released recently demonstrating that models with smaller sizes (4B and 27B) can achieve state-of-the-art performance in medical domain tasks including vision-language interactions. However, it is still poorly understood how different alignment stages—particularly CoT supervision and reward tuning—interact, or how alignment strategies scale across model sizes.

In this work, we focus on medically grounded QA tasks with known verifiable answers and introduce a modular framework for aligning small language models through staged CoT and GRPO tuning. We evaluate performance on both in-domain and out-of-domain datasets, and release a filtered, CoT-augmented dataset to support reproducible research in clinical alignment.

## 3 Dataset Overview

We use distinct datasets for supervised fine-tuning with Chain-of-Thought (SFT with CoT) and for Group Relative Policy Optimization (GRPO). For SFT with CoT, we use a mix of medical QA datasets with multiple-choice, short and long free-form answers. For all GRPO alignment configurations, we use a fixed datset with short free-form answers. Evaluation is performed on a held-out test set that combines three publicly available and common benchmark datasets—MedQA, OpenBookQA, and ARC Challenge—which are all multiple-choice Jin et al. [2021], Clark et al. [2018], Mihaylov et al. [2018].

### 3.1 SFT with CoT Training Datasets

#### FreedomIntelligence/medical-o1-reasoning-SFT

This dataset includes 19,704 medical QA samples with pre-generated CoT by GPT-4o and long multi-sentence free-form answers Chen et al. [2024]. We split this dataset 80/20 into 14,778 training and 4,926 validation samples.

#### FreedomIntelligence/medical-o1-verifiable-problem

Containing 40,644 medical QA samples with short phrase or single-sentence free-form answer Chen et al. [2024], this dataset was augmented with GPT-4o-generated CoT and filtered using LLM-as-judge evaluation (Mistral-3-24B). We retain only the samples that GPT-4o answered correctly, resulting in 14,880 training and 4,898 validation examples.

#### bigbio/med_qa

This dataset includes 12,723 multiple-choice medical questions with pre-defined training, validation, and test sets Jin et al. [2021]. CoT was generated with GPT-4o, and only examples where GPT-4o produced a correct answer (judged by Mistral-3-24B) in the training and validation set were retained: 8,884 for training and 1,110 for validation.

#### openlifescienceai/medmcqa

This dataset includes over 180K medical QA with multiple-choice answers Pal et al. [2022]. A subset of 59,994 training examples from the original MedMCQA dataset was used. CoT was generated with GPT-4o, and 51,020 examples with correct answers were selected for training.

#### lavita/AlpaCare-MedInstruct-52k

This dataset contains 52,002 long-form instruction-following QA samples Zhang et al. [2023b]. After CoT generation and filtering by correctness, we retain 28,809 training and 9,579 validation examples.

### 3.2 GRPO Training Dataset

For all GRPO training, we use the validation split of *FreedomIntelligence/medical-o1-verifiable-problem*, comprising 10,161 samples—note that this includes the questions that GPT-4o answered incorrectly that were excluded for SFT with CoT training. We fix the training set for GRPO for all alignment configurations since it allows for a consistent training signal across experiments. Other datasets such as MedQA, MedMCQA, and AlpaCare were excluded due to uncertainty in base model pretraining exposure or incompatibility with the GRPO reward structure.

### 3.3 Evaluation Datasets

All models are evaluated on the held-out test sets of:

- MedQA (medical domain)
- OpenBookQA (general science reasoning)
- ARC Challenge (general science reasoning)

For OpenBookQA, we evaluate without providing the supporting facts, treating it as a pure reasoning task. This allows us to test generalization and unintended degradation from medical domain-specific fine-tuning.

## 4 Methodology

### 4.1 Overview

We use a modular alignment framework that separates supervised reasoning and reward optimization into distinct parameter-efficient components. The approach is designed to support the training and evaluation of small language models in safety-critical domains such as medicine, where modularity, verifiability, and deployment constraints (e.g., privacy and limited compute) are central concerns.

Our training framework leverages Chain-of-Thought (CoT) supervision in the supervised fine-tuning (SFT) phase, followed by Group Relative Policy Optimization (GRPO) for aligning models to prefer verifiable, factual responses. Each stage is implemented using LoRA-based parameter-efficient fine-tuning (PEFT), enabling independent training and flexible composition of adapters. We compare multiple configurations of this pipeline to isolate the effects of CoT supervision, reward tuning, and adapter composition.

### 4.2 Alignment Configurations

We evaluate five model variants, each representing a different alignment strategy:

1. **Base (Zero-shot):** The original instruction-tuned model without any further fine-tuning.
2. **SFT Only:** A LoRA adapter fine-tuned with CoT-augmented supervised data. This model is optimized to generate reasoning traces and correct answers, but is not reward-aligned.
3. **GRPO Only:** A LoRA adapter trained via GRPO from the base model. This model receives no intermediate reasoning supervision and relies solely on reward feedback for alignment.
4. **Modular:** A two-stage configuration where a CoT-SFT adapter is frozen and a second, independent LoRA adapter is trained using GRPO. This setup enables modular reasoning and reward components to be combined at inference time.
5. **Unified** A single LoRA adapter trained sequentially with both CoT supervision and GRPO. This configuration reflects a tightly coupled alignment pipeline.

Each configuration is trained with the same initialization (from the base model) and evaluated using identical inference prompts and decoding parameters. We consistently use the following system message for all training and generation configurations:

You are a language model specialized in medical reasoning. Your task is to respond to queries by first thinking about the reasoning process in the mind and then providing the user with the answer. The reasoning process and answer are enclosed within <think> </think> and <answer> </answer> tags, respectively, i.e., <think> reasoning process here </think><answer> answer here </answer>

#### Models trained

We conduct experiments on the full range of Qwen2.5 0.5B–7B instruction fine-tuned models loaded with 4-bit quantization and fine-tuned using LoRA adapters for both supervised (CoT) and reward (GRPO) alignment phases.

#### Inference Settings

All model generations use consistent system message, prompt for each question, and decoding parameters across SFT, GRPO, or base instruction fine-tuned models:

- Temperature: 0.0 and Top-*p*: 1.0
- Max prompt length: 1024 tokens, Max completion length: 1024 tokens

#### Compute resources

For all model trainings, we use 1-2 NVIDIA H100 GPUs on a Debian Linux system with 1TB of RAM. PyTorch and HuggingFace was extensively used for all development Paszke et al. [2019], Wolf et al. [2020].

### 4.3 Parameter-Efficient Fine-Tuning (PEFT)

We use LoRA adapters for all SFT and GRPO stages Hu et al. [2022]. All adapters are trained with consistent hyperparameters: *r* = 16, *α* = 32, dropout = 0.05 and applied to the following target modules `[q_proj, k_proj, v_proj, o_proj, up_proj, down_proj, gate_proj]`.

### 4.4 Chain-of-Thought (CoT) Supervision

In the SFT phase, models are trained using multiple-choice, short, and long free-from medical QA data augmented with CoT reasoning traces generated by GPT-4o. We filter training examples using LLM-as-judge verification (Mistral-3-24B) and retain only those for which GPT-4o answers correctly. Each example is formatted as a chat-like conversation:

- **System message:** as provided above
- **User message:** a medical question, either as a sentence free-form query or with multiple choice options in the format `A. Option 1, B. Option 2`, etc. depending on the dataset.
- **Assistant message:** a step-by-step explanation followed by a final answer using the `<think></think> and <answer></answer>` format.

The model is trained to reproduce both the reasoning trace and answer. The complete set of COT SFT training parameters are provided in Appendix.

### 4.5 Group Relative Policy Optimization (GRPO)

We apply GRPO to align models with verifiability and factuality without requiring human preference labels. We use the same system message as described previously. Additionally, the training setup includes:

- **User message:** a medical question as a sentence free-form query
- **Candidate generations:** sampled from the model with diversity using temperature sampling
- **Reward model:** we combine a format reward and an accuracy reward as done in other work

#### Algorithm 1

Accuracy Reward with Text Matching

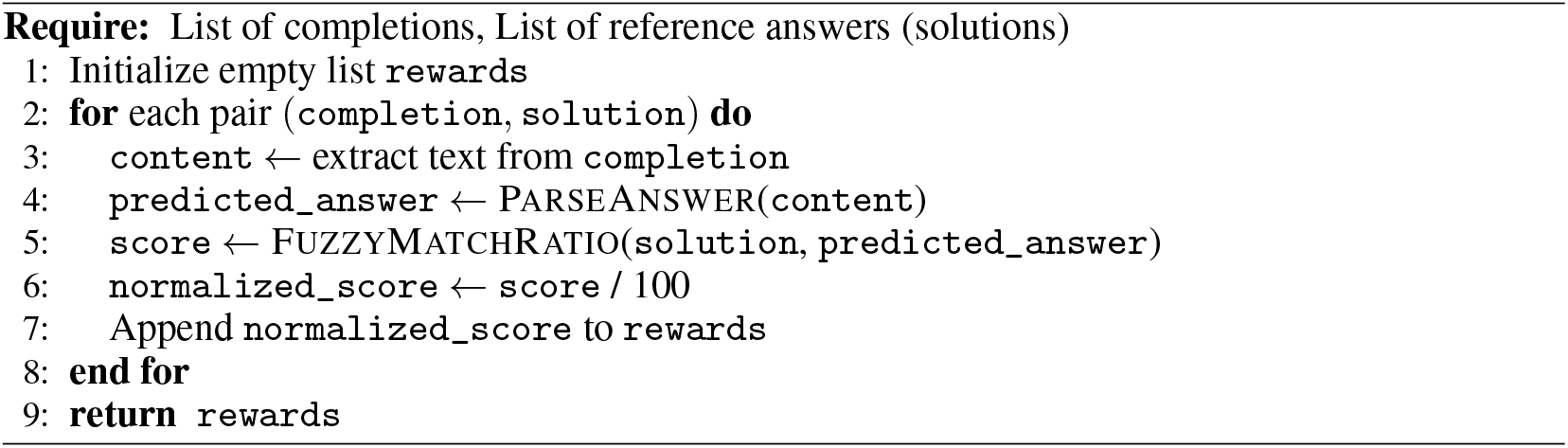

#### Algorithm 2

ParseAnswer Function

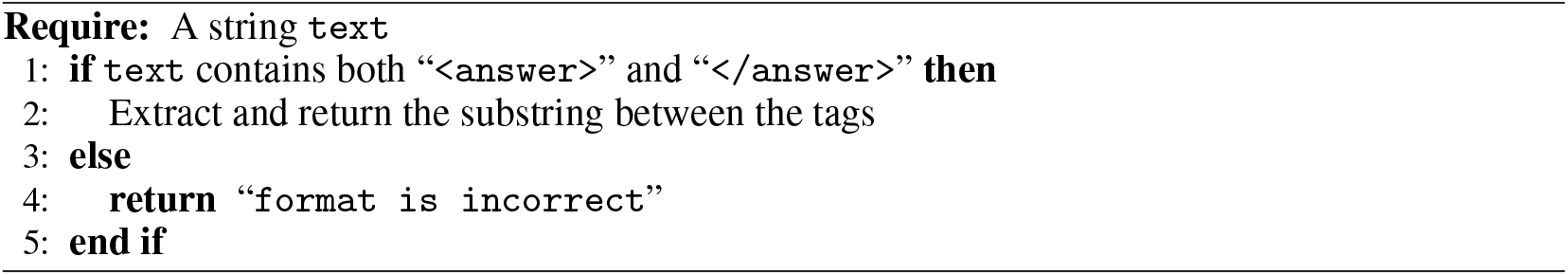

- **Preference pairs:** automatically ranked pairs where the preferred output receives a higher reward

GRPO is applied to either a fresh LoRA adapter (GRPO only and modular setups) or directly to the SFT adapter (unified setup). The complete set of GRPO training parameters are provided in Appendix. The reward functions are described further below.

#### 4.5.1 GRPO reward models

We sum outputs from *format reward* and *accuracy reward* functions to assign a reward to each generation Shao et al. [2024], Guo et al. [2025]. Each reward function can return a minimum value of 0 and a maximum value of 1. The format reward is unchanged from previous work but the accuracy reward is modified to the specific problem.

The *format reward* assigns 1.0 if the model’output adheres to a specific structure, where reasoning text is enclosed in `<think>` tags and the answer text in `<answer>` tags. Otherwise, it returns 0.0.

The *accuracy reward* is computed by comparing the predicted answer text (candidate generations) to a reference solution (ground truth answer only, no CoT supervision) using fuzzy string matching (refer to Algorithms 1 and 2). Each model completion is expected to follow a specific structured format, where the final answer is enclosed in `<answer>` tags. The function first parses the output to extract the answer segment, then uses the Levenshtein ratio (via fuzzy matching) to compute a similarity score between the model’s predicted answer and the gold answer. This score is normalized to fall between 0 and 1, and serves as the accuracy reward signal.

This approach provides a soft reward that tolerates minor variations in phrasing, while still encouraging exactness in the model’s final answer.

### 4.6 Evaluation Metrics

We report two key metrics for model comparison:

#### Format Correctness

We compute the proportion of responses that are in the expected format with `<think></think>` and `<answer></answer>` text containing reasoning and the answer.

#### Answer Accuracy

We parse any string contained within the first `<answer></answer>` for all responses and compute the proportion of matches between the model’s final answer and the gold label using LLM-as-judge (Mistral-3-24B). Please note that the *format may be incorrect overall but the answer may be judged to be accurate* since it was embedded within the `<answer></answer>` tags.

## 5 Results

### 5.1 GRPO training dynamics

We visualize the total reward (format + accuracy) during GRPO training across the first 1000 steps for Qwen2.5 models at four scales: 0.5B, 1.5B, 3B, and 7B (Figure 1).

**Figure 1.**
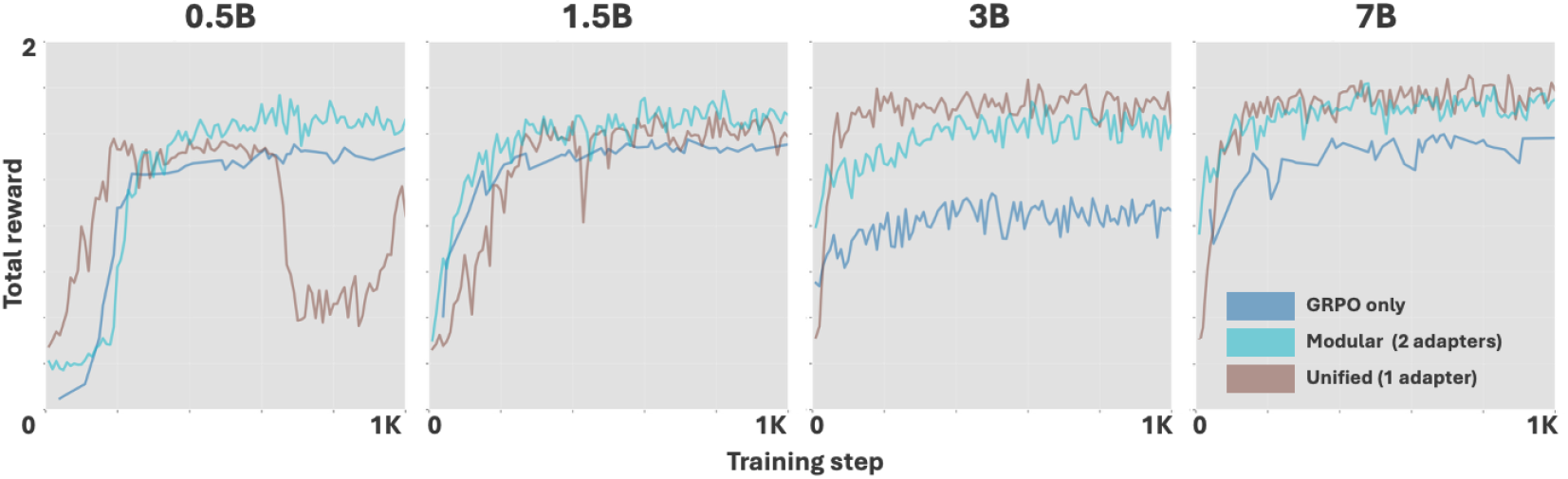
GRPO training dynamics across Qwen2.5 model scales (0.5B to 7B) under 3 alignment configurations: GRPO only (blue), Modular (cyan), and Unified (brown), for the first 1000 steps. While smaller models (e.g., 0.5B) exhibit less stability—especially for the *unified* adapter setup—larger models converge more smoothly. The *modular* configuration consistently achieves higher total reward with greater stability across scales.

#### Convergence and Stability

We observe that all models converge to high reward levels within the first 500–800 steps, consistent with findings from prior work on reward preference optimization. However, training dynamics vary significantly by model size and configuration.

#### Small model instability

For the 0.5B model, the *unified* setup (shared adapter for SFT and GRPO) initially achieves the fastest reward gain but exhibits training collapse between steps 500–900, suggesting interference between reasoning and reward objectives when trained in a single adapter. In contrast, the *modular* setup (independent adapters for SFT and GRPO) proves resilient to this collapse, achieving higher final reward with greater stability.

#### Consistent modular advantage

The *modular* configuration consistently outperforms the *GRPO only* baseline in all models and achieves the highest final reward in the 0.5B, 1.5B, and 7B models. This suggests that pretraining with CoT reasoning provides a robust foundation that accelerates reward optimization.

#### Unified vs. Modular trade-offs

In the 3B and 7B models, the *unified* configuration performs comparably to the *modular* approach, with slightly higher peak reward in the 3B setting. This indicates that larger models may be more robust to multi-objective training in a single adapter, while smaller models benefit from explicit modularization.

### 5.2 Format correctness

We assess models’ ability to produce outputs in the expected reasoning format—specifically, generating step-by-step reasoning within `<think>` tags followed by a final answer enclosed in `<answer>` tags. This structural fidelity is essential in clinical and safety-critical contexts, where verifiable formatting supports downstream auditability and response validation. Figure 2 (top row) shows format correctness across models, datasets, and alignment configurations.

**Figure 2.**
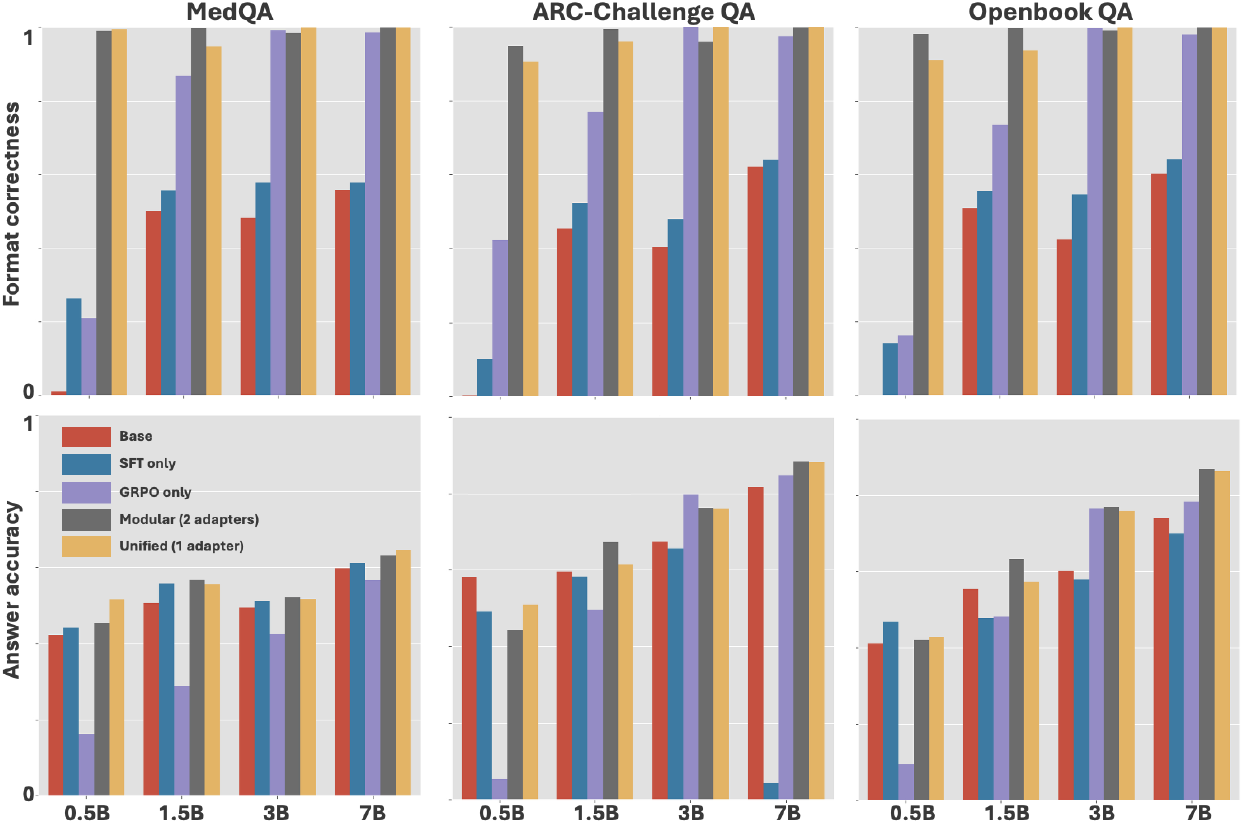
Format correctness (top row) and answer accuracy (bottom row) across model sizes (0.5B–7B) and alignment configurations on MedQA, ARC-Challenge, and OpenBookQA. Modular alignment (gray) consistently achieves the highest format correctness across all datasets, especially at smaller model scales. While Unified alignment (gold) also yields strong structure adherence, its accuracy can lag slightly behind the modular variant. GRPO only models benefit from reward tuning but often fail to consistently produce outputs in the correct reasoning format, particularly at smaller scales. Larger models (e.g., 7B) show less variance across configurations, suggesting diminishing returns from modularity at higher capacities. Note that the answer can be accurate without the overall format being correct since we evaluate any text within `<answer></answer>` tags as the answer.

### Modular alignment promotes structural adherence

*Modular* configuration consistently achieves high format correctness across all datasets and model sizes. For example, even the 0.5B model maintains near-perfect format correctness, a notable contrast to the *baseline, SFT only and GRPO only* variants. This indicates that decoupling reasoning supervision and reward tuning into separate adapters leads to clean and structurally consistent outputs which are a critical requirement for building auditable and verifiable clinical systems Garg et al. [2025], Jonnagaddala and Wong [2025].

#### Unified alignment performs less consistently in smaller models

The *unified* configuration also yields high format adherence, often comparable to the *modular* setup at larger model scales. However, its performance is less consistent at smaller sizes. For instance, the 0.5B and 1.5B models exhibit mild degradation in format consistency, particularly in ARC and OpenBookQA, suggesting that training multiple objectives in a single adapter can introduce instability in smaller models.

#### Effect of model scale

Larger models (3B and 7B) exhibit high format correctness across all GRPO configurations, suggesting that model scale can enforce format adherence with *GRPO only* and no prior COT supervision. On the other hand, *SFT only* consistently fails to follow the prescribed output structure across all model sizes, indicating that reward-alignment is crucial to instill structural consistency.

### 5.3 Answer accuracy

To assess the impact of different alignment strategies on factual correctness, we evaluate answer accuracy across three benchmarks—MedQA, ARC-Challenge, and OpenBookQA—using four Qwen2.5 model sizes (0.5B, 1.5B, 3B, 7B). The bottom row of Figure 2 shows the accuracy results for each combination.

#### Effect of Model Scale

Unsurprisingly, answer accuracy improves consistently with model size across all configurations and datasets. The 7B models outperform smaller models by a large margin, particularly on ARC-Challenge and OpenBookQA, where larger-scale LLMs demonstrate stronger generalization to science reasoning tasks. Interestingly, even for 0.5B and 1.5B models, *modular* alignment strategy significantly improves accuracy relative to the base model, suggesting that small models can still benefit from decoupled fine-tuning even in challenging tasks.

#### Modular vs. Unified Alignment

The best-performing models across nearly all settings are those trained with both SFT and GRPO. Notably, the *modular* alignment strategy tends to outperform the *unified* single-adapter configuration in some model sizes and datasets. This suggests that isolating reasoning supervision from reward objectives can allow for more stable optimization and improved factual correctness, especially in smaller models (e.g., 0.5B and 1.5B). In the 7B models, both multi-stage configurations achieve similar accuracy, implying that larger models may internally disentangle reasoning and reward even when co-trained in a unified adapter.

#### Base vs. SFT

Models fine-tuned with CoT supervision (*SFT only*) outperform the base model in MedQA dataset, demonstrating the benefit of reasoning traces in enhancing factual grounding for in-domain datasets. In contrast, the performance degrades in science reasoning tasks indicating poor generalization to out-of-domain datasets.

#### GRPO Only

Large Models (3B and 7B) trained only with GRPO reward tuning achieve surpassing the base and *SFT only* configurations on out-of-domain science reasoning tasks. This highlights the value of reward tuning for large models, even when not scaffolded by reasoning supervision.

## 6 Discussion and Limitations

Our experiments demonstrate that *modular* alignment using separate adapters for Chain-of-Thought (CoT) supervision and reward optimization consistently improves model performance across scale, structure adherence, and factual accuracy. These findings offer a path forward in resolving the tension between accuracy, auditability, and efficiency in clinical LLMs. Our work highlights a significant pattern: applying reward optimization like GRPO without a strong foundation of supervised, structured reasoning can lead to illusory performance, or even degrade in-domain accuracy, especially in smaller models.

### Modular GRPO Training Improves Stability

Training reward adapters separately from reasoning adapters leads to more stable optimization and higher final reward, particularly for smaller models. *Unified* (single adapter) setups perform comparably at larger scales but show instability in 0.5B and 1.5B models—maybe due to conflicting objectives.

### Structural Format Adherence is Best with Modular Setup

The GRPO only configuration— trained without CoT traces—frequently fails to follow the required output structure. *SFT only* models follow the format reliably, but lack reinforcement to consistently maintain structure in out-of-domain settings. *Unified* models perform well structurally at larger scales, but degrade under 3B.

### Answer Accuracy Benefits from Modular Alignment

Models trained with both SFT and GRPO yield the best factual performance, especially under the *modular* configuration. This advantage is clearest for small and mid-sized models, while 7B models benefit less from decoupling—suggesting that larger models may inherently disentangle alignment objectives. This modularity can offer a practical advantage. As clinical guidelines evolve and new knowledge emerges, with separate adapters, a clinical institution could rapidly update or retrain a ‘reward’ adapter to align with a new standard of care without needing to retrain the foundational reasoning adapter.

### Dataset-Specific Trends

GRPO only models do not improve on MedQA, suggesting that factual grounding alone is insufficient without explicit reasoning supervision. In contrast, SFT-trained models (with or without GRPO) consistently improve on MedQA. For ARC-Challenge and OpenBookQA, GRPO-based alignment reliably boosts accuracy, particularly in the 3B and 7B settings—indicating that reward tuning generalizes well beyond the medical domain when scaffolded by CoT.

### Dependency on COT Traces

While the *modular* alignment framework improves stability and factual correctness, it is limited by the availability of clean COT traces for initial alignment. In real-world or domain-specific settings, such traces may be scarce and difficult to collect at scale. In practice, this often necessitates the use of LLMs to generate traces which can introduce clinical inaccuracies, ulitmately reducing the reliability of downstream alignment.

### Training and Scalability Trade-off

*Unified and modular* setups introduce an additional fine-tuning stage for supervised CoT reasoning and increase the total number of trainable parameters. This overhead is particularly valuable to stabilize GRPO training in smaller models. However, as gains become more marginal with larger models, this setup may demand higher resources as datasets and models scale.

Overall, our findings highlight the practical value of *modular* alignment pipelines—particularly for small, efficient models intended for use in verifiable or safety-critical domains.

## Data Availability

All data used is Public available at Hugging face repository

## A Appendix

### A.1 Human validation of LLM-as-judge

We performed human evaluation for 100 generated responses to validate the LLM-as-judge since all evaluation metrics depend upon the validity of LLM-as-judge. We psuedo-randomly selected 50 responses where the LLM-as-judge determined that the generated response was incorrect and 50 where it determined the response was correct. The human was asked to determine whether the generated response aligned with the ground truth answer. The human was blinded to the decision of LLM-as-judge.

We find total agreement between the human determination and LLM-as-judge for the samples selected for human evaluation.

**Table 1:** Confusion matrix comparing human evaluation and LLM-as-judge labels for 100 QA pairs.

| Human Label | LLM: Correct | LLM: Incorrect |
| --- | --- | --- |
| Correct | TP (50) | FN (0) |
| Incorrect | FP (0) | TN (50) |

### A.2 Chain-of-Thought Supervised Fine-Tuning Hyperparameters (CoT SFT)

| *Optimization* |  | *Training Setup* |  |
| --- | --- | --- | --- |
| Learning rate | $2 \times 10^{-4}$ | Batch size | Dynamic |
| Optimizer | AdamW (8-bit) | Epochs | 2 |
| Adam $(\beta_1, \beta_2)$ | (0.9, 0.999) | Grad. acc. steps | 4 |
| Adam $\epsilon$ | $1 \times 10^{-8}$ | Max grad norm | 1.0 |
| Weight decay | 0.01 | Scheduler | Cosine |
| Warmup steps | 100 |  |  |
| *Input Configuration* |  | *Model Context* |  |
| Max input length | 1024 tokens | Dataset field | text |

### A.3 GRPO Training HyperParameters

| *Optimization* |  | *Training Setup* |  |
| --- | --- | --- | --- |
| Learning rate | $1 \times 10^{-5}$ | Batch size | 8 / 8 |
| Optimizer | AdamW | Epochs | 2 |
| Adam $(\beta_1, \beta_2)$ | (0.9, 0.999) | Grad. acc. steps | 8 |
| Adam $\epsilon$ | $1 \times 10^{-8}$ | Max grad norm | 1.0 |
| Weight decay | 0.0 | Loss type | BNPO |
| *Generation Parameters* |  | *Precision and Device* |  |
| Temp. / Top-$p$ / Top-$k$ | 0.9 / 1.0 / 50 | FP16 / BF16 | On / Off |
| Prompt / Completion length | 1024 / 256 | CUDA enabled | True |
| Completions per prompt | 64 | Repetition penalty | 1.0 |
| *Reward Configuration* |  |  |  |
| Reward type | Format + Fuzzy Match | Reward scaling | Enabled |
| Epsilon (clip) | 0.2 | Ref. mixup $\alpha$ | 0.6 |
| Ref. sync steps | 512 |  |  |

